# Regional Disparities in Undiagnosed Hypertension Among South Asian Adults in the Bay Area and Central Valley: The PRANA Heart Study

**DOI:** 10.64898/2026.09.13.26362810

**Authors:** Vivek Nalluri, Dilsi Bhagat, Shreyaa Gunasekar, Billy Zeng, Seema Policepatil, Winston Tseng

## Abstract

**Background:** South Asians (from Bangladesh, Bhutan, India, the Maldives, Nepal, Pakistan, and Sri Lanka) experience a disproportionately high burden of cardiovascular disease, with hypertension serving as a major modifiable risk factor. Despite this elevated risk, hypertension often remains underdiagnosed, particularly in community settings with limited access to routine screening. This study assessed the prevalence of hypertension and undiagnosed hypertension among South Asian adults participating in community-based health screenings in Northern California and examined regional disparities between the San Francisco Bay Area and the Central Valley.

**Methods:** A cross-sectional survey-based study was conducted among adults attending screenings organized by Jeeva Clinic at religious and community institutions. Participants voluntarily completed a survey collecting demographic information, cardiovascular health history, lifestyle factors, and recent blood pressure measurements obtained either during screening or through self-report. A total of 300 participants were included in the final analysis.

**Results:** Among participants with sufficient blood pressure data for hypertension classification (n = 245), 82.9% met criteria for hypertension. Among participants with sufficient data to determine undiagnosed hypertension status (n = 244), 59.8% exhibited previously undiagnosed hypertension. Hypertension prevalence was significantly higher among participants in the Central Valley compared with the Bay Area (97.9% vs 71.0%, χ² = 31.2, p < 0.001). Undiagnosed hypertension was also substantially more prevalent in the Central Valley (91.7% vs 37.0%, χ² = 71.0, p < 0.001). In multivariable analysis, Central Valley region was associated with significantly higher odds of hypertension (OR = 14.29, 95% CI 2.89–70.63, p = 0.001), and higher BMI was independently associated with hypertension (OR = 1.26 per unit increase, 95% CI 1.10–1.43, p < 0.001).

**Conclusions:** These findings highlight a high burden of hypertension and significant regional disparities among South Asian adults in Northern California, suggesting that a large proportion of cardiovascular risk in this population remains undetected in routine care settings. Community-based screening initiatives, such as Jeeva Clinic, may help identify individuals at elevated risk and improve early detection in underserved populations.

## Introduction

Cardiovascular disease (CVD) remains the leading cause of mortality worldwide and is a major contributor to morbidity in the United States.^1, 2^ Hypertension is one of the most significant modifiable risk factors for CVD and is strongly associated with stroke, myocardial infarction, and heart failure.^3^ Early detection and management of hypertension are therefore critical components of cardiovascular disease prevention.

South Asian populations experience a disproportionately high burden of cardiovascular disease compared with many other ethnic groups.^4–6, 12^ Studies have shown that individuals of South Asian ancestry develop cardiovascular disease at younger ages and often at lower body mass index levels than other populations.^5, 7, 13^ In addition to genetic predisposition, lifestyle factors and structural barriers to preventive care may contribute to elevated cardiovascular risk within these communities.^6, 8^

Despite the well-documented cardiovascular risk among South Asians, hypertension often remains underdiagnosed and undertreated, particularly in community settings where routine screening may be limited.^9^ Regional disparities in healthcare access may further exacerbate this issue. The Central Valley of California has historically experienced lower healthcare access and higher chronic disease burden compared with urban regions such as the San Francisco Bay Area.^10,11^ Understanding patterns of hypertension prevalence and diagnosis across these regions may provide insight into potential gaps in cardiovascular screening and care.

The present study aimed to assess the prevalence of hypertension and other cardiovascular risk factors among South Asian adults participating in community health screenings in Northern California. In addition, we sought to evaluate the prevalence of previously undiagnosed hypertension and examine regional differences between participants residing in the Bay Area and those residing in the Central Valley.

## Methods

### Study Design and Setting

This study was a cross-sectional survey-based analysis of cardiovascular risk factors among adults participating in community health screenings organized by Jeeva Clinic.^14^ Jeeva Clinic is a student-led organization that conducts screenings at religious and community institutions, including temples, churches, and mosques, across Northern California, with a primary focus on the San Francisco Bay Area. These screening events are part of Jeeva Clinic’s ongoing outreach efforts aimed at improving cardiovascular health awareness and expanding access to preventive screening services in underserved communities. Community-based screening approaches have been increasingly utilized to improve preventive health engagement in underserved populations.^15^

The screening events primarily served South Asian populations, including individuals of Indian, Pakistani, Bangladeshi, and Nepali descent, a group known to be at elevated risk for cardiometabolic disease.

Participants attending these events were invited to voluntarily complete a research survey collecting demographic information, cardiovascular health history, and lifestyle factors. Participation was optional and did not affect access to screening services.

### Participants and Data Collection

Adults (≥18 years) attending Jeeva Clinic screening events were eligible to participate in the study. Individuals who chose to participate completed a survey collecting information on demographics, cardiovascular health history, lifestyle behaviors, and recent blood pressure measurements.

Blood pressure values were obtained either from measurements conducted during the Jeeva Clinic screening event or from participants’ self-reported most recent blood pressure readings if measurements were not performed at the event.^16, 17^ Because blood pressure data were collected in both measured and self-reported formats, the dataset reflects a combination of direct clinical measurements and participant-reported values.

Participants were also asked to report prior diagnoses of hypertension, diabetes, high cholesterol, and heart disease. Additional variables collected included exercise frequency (days per week), sleep duration (hours per night), perceived stress levels (five-point scale), and healthcare utilization patterns.

Data collection occurred across multiple community screening events, with the final day of data collection on February 22, 2026. A total of 319 survey responses were collected prior to data cleaning.

### Geographic Classification

Participants were categorized into geographic regions based on self-reported county of residence. In the final dataset, participants from the San Francisco Bay Area were primarily from Alameda, Contra Costa, San Mateo, and Santa Clara counties, while participants from the Central Valley were from San Joaquin and Merced counties. Participants reporting residence outside these regions (e.g., Southern California or outside the United States) were excluded from regional comparative analyses.

### Data Cleaning and Variable Construction

Survey responses were reviewed and cleaned prior to analysis to ensure data completeness and internal consistency. Duplicate responses were excluded based on participant self-report. Responses with missing or “don’t remember” values were retained in the overall analytical dataset when sufficient data were available for other analyses. Analyses requiring specific variables were conducted using available-case data, with participants missing the variables required for a given analysis excluded from that analysis. After data cleaning, 300 responses were included in the final analytical dataset.

Continuous variables such as height and weight were standardized by converting reported units (feet/inches to centimeters; pounds to kilograms), and body mass index (BMI) was calculated as weight (kg) divided by height squared (m²). Implausible anthropometric values were excluded.

Blood pressure values were collected as categorical ranges for some participants. To enable descriptive analysis of mean blood pressure, categorical responses were converted to midpoint estimates. For example, systolic categories of 120–129 mmHg and 130–139 mmHg were assigned midpoint values of 125 mmHg and 135 mmHg, respectively. Open-ended categories (e.g., “<120” or “≥180”) were assigned reasonable midpoint approximations based on standard clinical thresholds.

### Objectives

Our primary objective was to look at the prevalence of hypertension. Routine blood pressure screening is recommended for early identification of hypertension in adult populations.^16^ Blood pressure categories were defined according to the 2017 American College of Cardiology/American Heart Association (ACC/AHA) guidelines: normal (<120/<80 mmHg), elevated (120–129/<80 mmHg), Stage 1 hypertension (130–139 mmHg or 80–89 mmHg), and Stage 2 hypertension (≥140 mmHg or ≥90 mmHg). Participants were classified as hypertensive if they met criteria for Stage 1 or Stage 2 hypertension.^18^ Hypertension status was determined only for participants with sufficient systolic and diastolic blood pressure data to establish an ACC/AHA blood pressure classification.

Undiagnosed hypertension was defined as participants who met criteria for hypertension based on measured or reported blood pressure values but did not report a prior diagnosis of hypertension on the survey. Participants who indicated a prior diagnosis of hypertension were classified as diagnosed cases regardless of current blood pressure values.

Secondary outcomes included the prevalence of diabetes, high cholesterol, and heart disease, as well as associations between hypertension and lifestyle and anthropometric factors including BMI, exercise frequency, sleep duration, and perceived stress.

### Statistical Analysis

Descriptive statistics were used to summarize participant demographics and cardiovascular risk factors. Categorical variables were reported as counts and percentages, while continuous variables were reported as means with standard deviations.

Differences in anthropometric and lifestyle variables between participants with and without hypertension were assessed using independent samples t-tests. Regional differences in hypertension prevalence and undiagnosed hypertension were evaluated using chi-square tests across geographic regions.

To further evaluate factors associated with hypertension, multivariable logistic regression was performed, adjusting for region, age group, gender, and BMI. Results were reported as odds ratios (ORs) with 95% confidence intervals (CIs).

Analyses were conducted using available cases for the variables included in each analysis. All analyses were conducted using jamovi (version 2.7).^37^ Statistical significance was defined as p < 0.05.

### Ethics Approval and Consent to Participate

This study was approved by the University of California, Berkeley Institutional Review Board (IRB Protocol #: 2025-10-19092). Participation in the survey was voluntary and anonymous. Participants were informed about the purpose of the study and provided informed consent prior to completing the survey. Participation or non-participation did not affect access to Jeeva Clinic screening services.

## Results

A total of **300 participants** were included in the analysis. Among participants with available ethnicity data (n = 298), the sample was **predominantly Indian (96.0%)**, with smaller proportions identifying as Pakistani (2.3%), Bangladeshi (1.0%), or Nepali (0.7%). Participants were distributed across age groups, with the largest proportion aged **45-54 years (30.7%)**, followed by **35-44 years (27.0%)** and **25-34 years (21.3%)**. The sample was relatively balanced by gender, with **156 male participants (52.0%)** and **144 female participants (48.0%)**. Most participants were from the **Bay Area (61.7%)**, while **33.0% were from the Central Valley** and **5.3% from other regions**.

Mean **BMI was 27.2 ± 3.73 kg/m²**. The mean systolic and diastolic blood pressures were **137 ± 17.7 mmHg and 81.4 ± 10.5 mmHg**, respectively. Mean **sleep duration was 6.41 ± 1.31 hours per night**, and participants reported exercising **2.65 ± 1.62 days per week**. The average self-reported stress score was **3.46 ± 1.09** on a five-point scale.

**Table 1.**
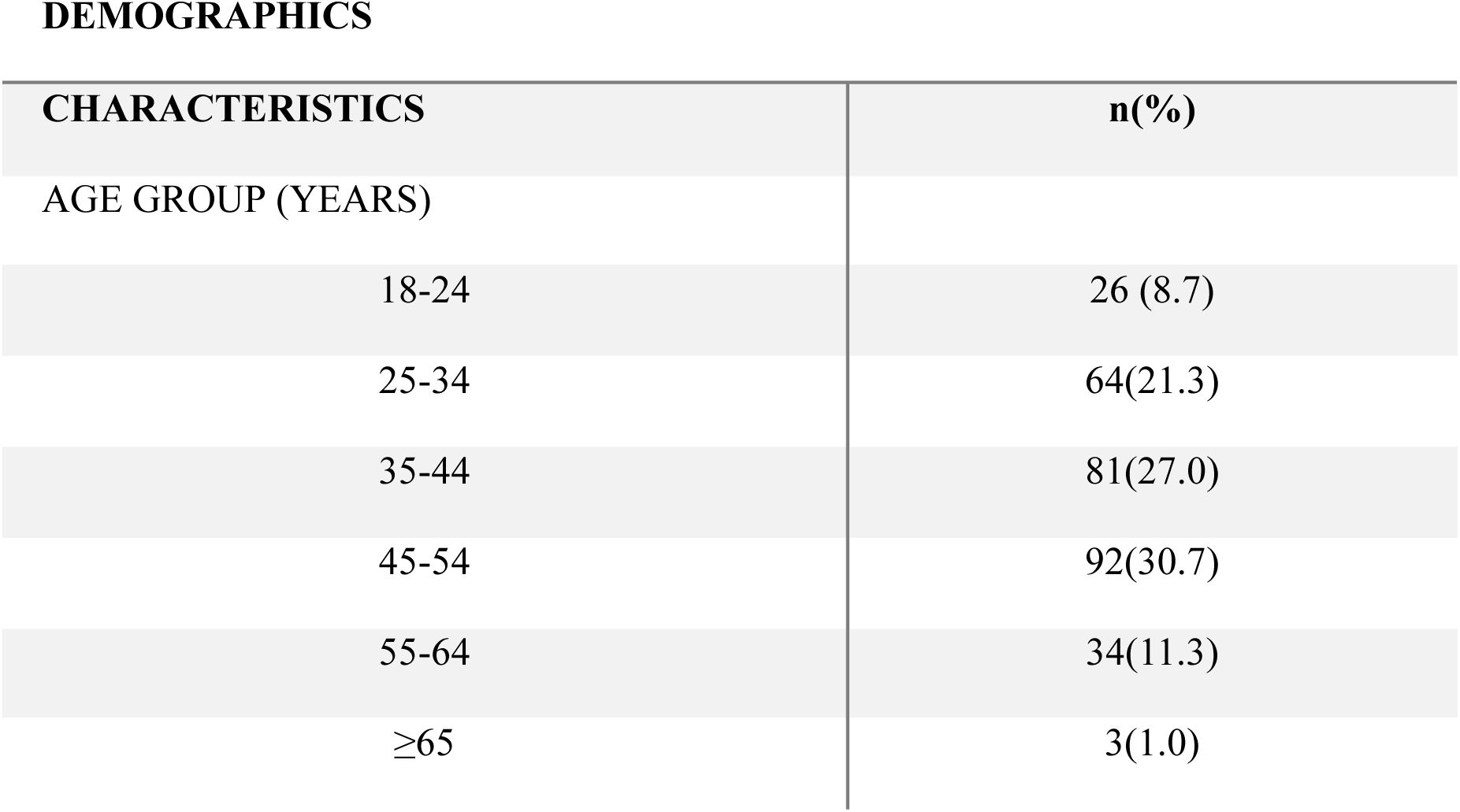
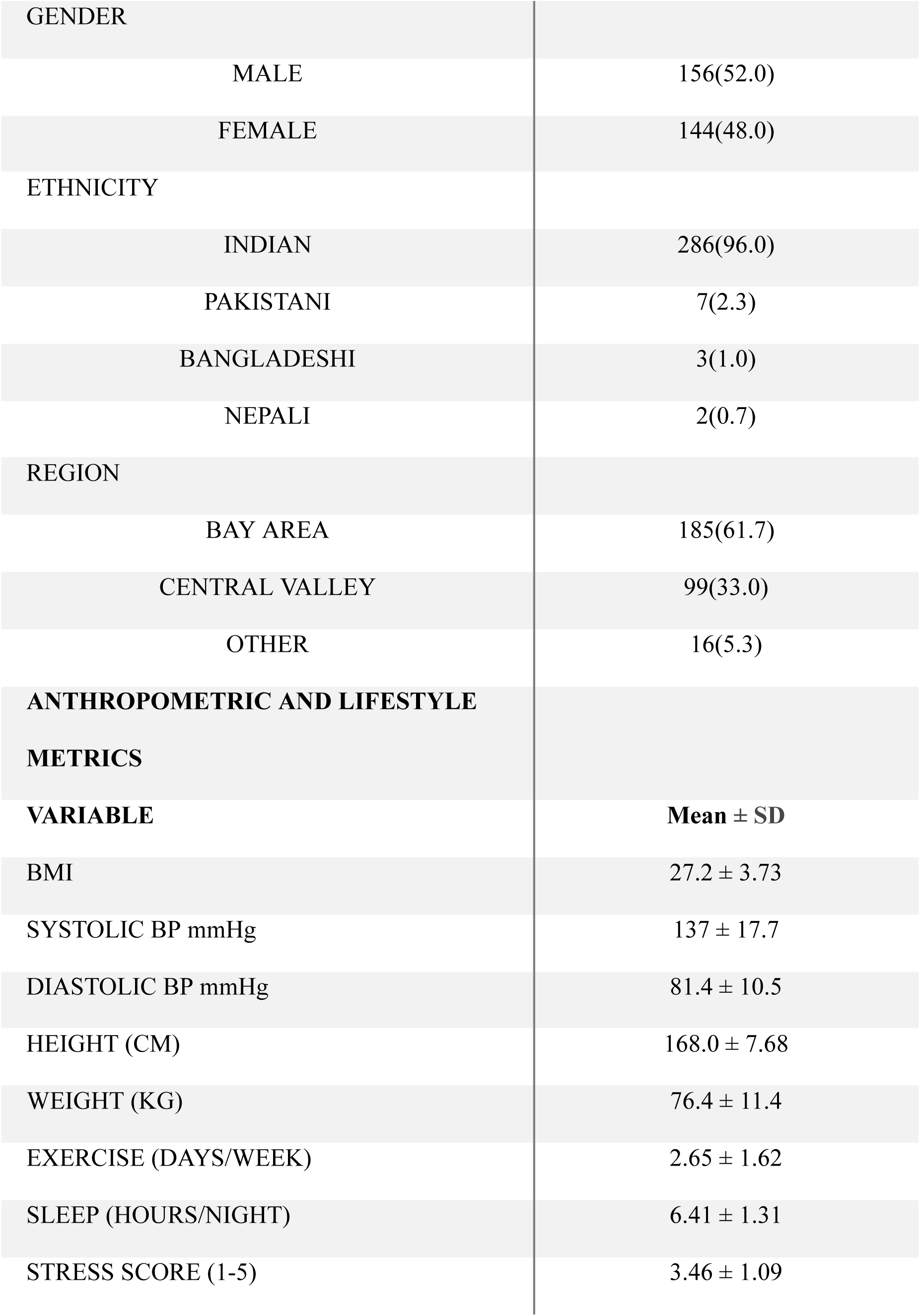
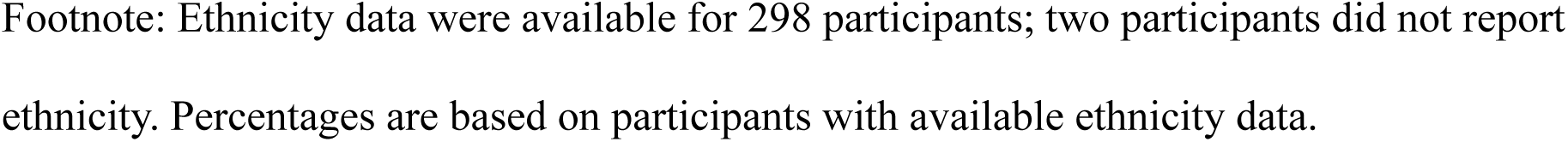
Participant Characteristics (N = 300)

Cardiovascular risk factors were common among participants. Among participants with sufficient blood pressure data for hypertension classification **(n = 245), 203 participants (82.9%) met criteria for hypertension**. Only **62 of 287 participants with available diagnosis history (21.6%) reported a prior diagnosis of hypertension**, while **146 of 244 participants with sufficient data to determine undiagnosed hypertension status (59.8%) met criteria for hypertension without reporting a previous diagnosis**.

Other cardiometabolic conditions were less common. **Diabetes was reported by 34 participants (11.8%)**, **high cholesterol by 45 participants (15.7%)**, and **heart disease by 5 participants (1.7%)**. Blood pressure classification revealed that **51.8% of participants were in Stage 2 hypertension**, while **31.0% were classified as Stage 1 hypertension**.

**Table 2.**
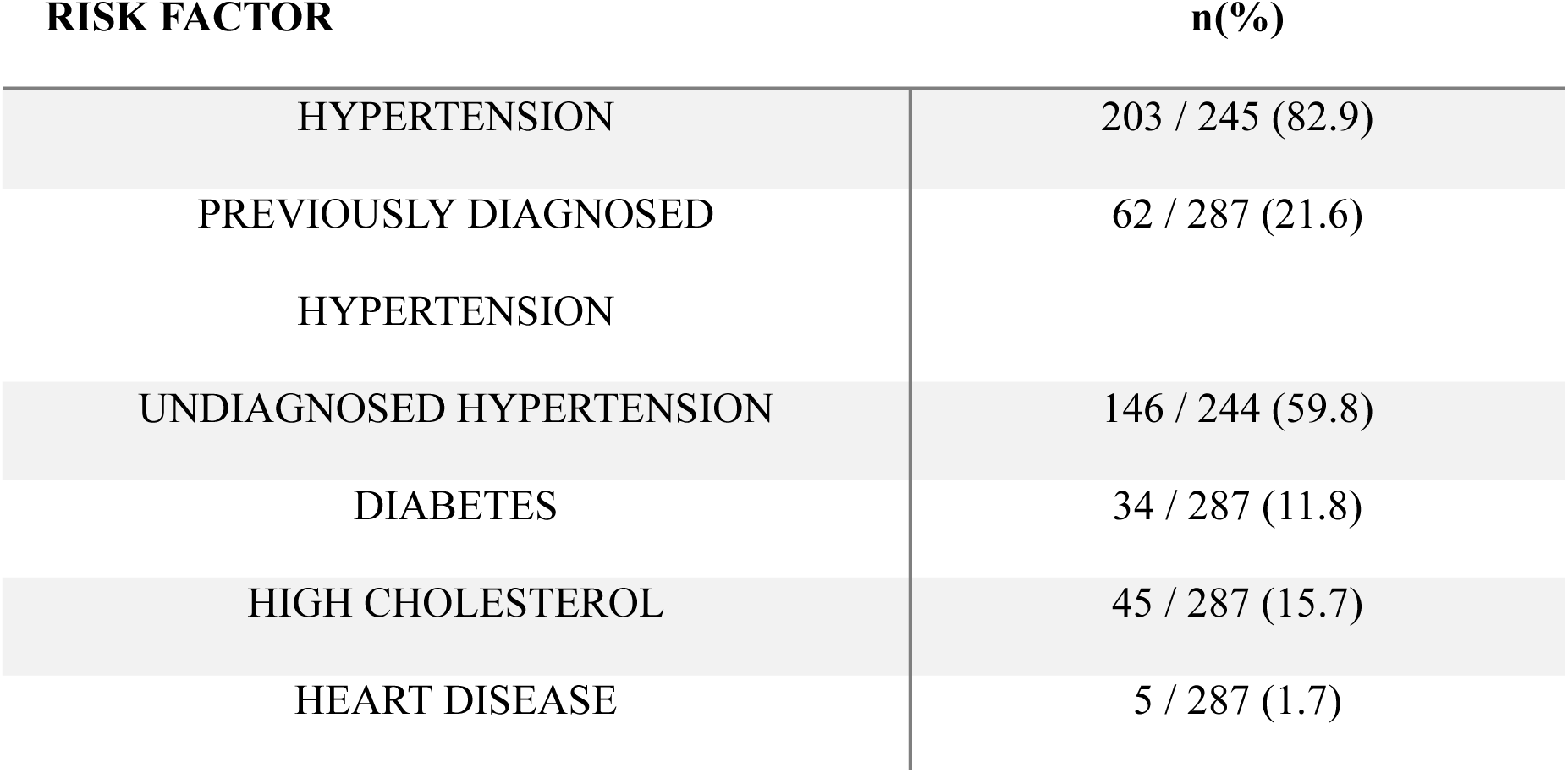
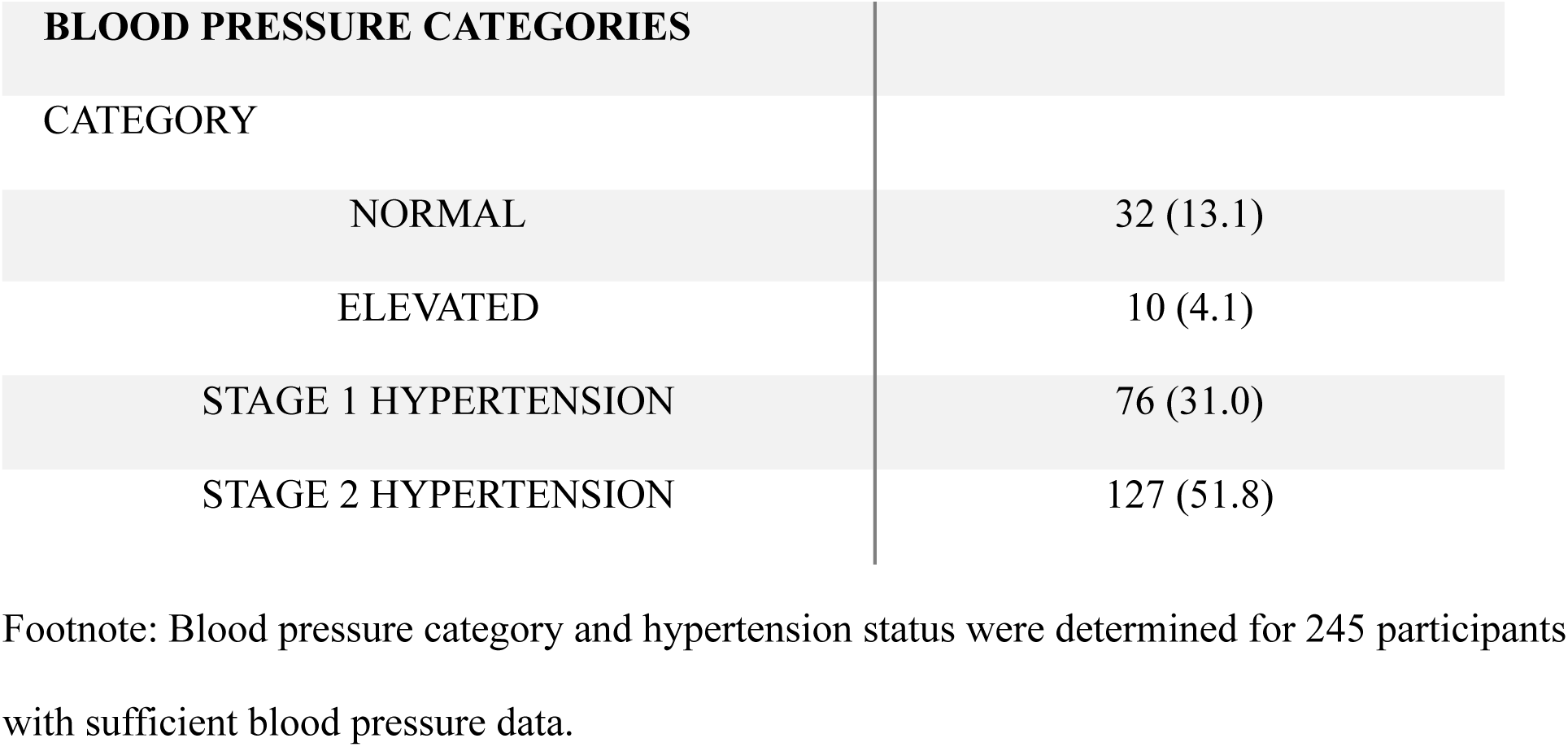
Prevalence of Cardiovascular Risk Factors.

| **RISK FACTOR** | **n(%)** |
| --- | --- |
| HYPERTENSION | 203 / 245 (82.9) |
| PREVIOUSLY DIAGNOSED<br>HYPERTENSION | 62 / 287 (21.6) |
| UNDIAGNOSED HYPERTENSION | 146 / 244 (59.8) |
| DIABETES | 34 / 287 (11.8) |
| HIGH CHOLESTEROL | 45 / 287 (15.7) |
| HEART DISEASE | 5 / 287 (1.7) |

| BLOOD PRESSURE CATEGORIES |  |
| --- | --- |
| CATEGORY |  |
| NORMAL | 32 (13.1) |
| ELEVATED | 10 (4.1) |
| STAGE 1 HYPERTENSION | 76 (31.0) |
| STAGE 2 HYPERTENSION | 127 (51.8) |

Footnote: Blood pressure category and hypertension status were determined for 245 participants with sufficient blood pressure data.

Participants with hypertension differed significantly from those without hypertension across several lifestyle and anthropometric measures. Individuals with hypertension had significantly **higher BMI** compared with participants without hypertension **(28.19 ± 3.43 vs 24.39 ± 2.70**, p < 0.001). Participants without hypertension reported **greater exercise frequency (3.06 ± 1.59 vs 2.46 ± 1.64 days per week**, p = 0.031). Sleep duration did not differ significantly between participants without and with hypertension (6.63 ± 1.18 vs 6.26 ± 1.35 hours per night, p = 0.103). Participants with hypertension reported significantly **higher stress scores (3.67 ± 1.07 vs 3.00 ± 1.01**, p < 0.001).

**Table 3.**
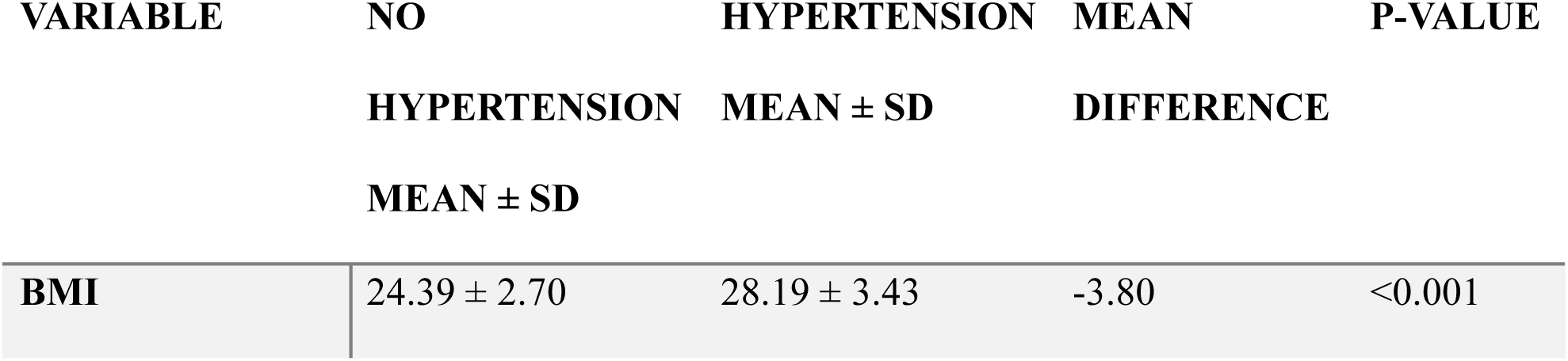
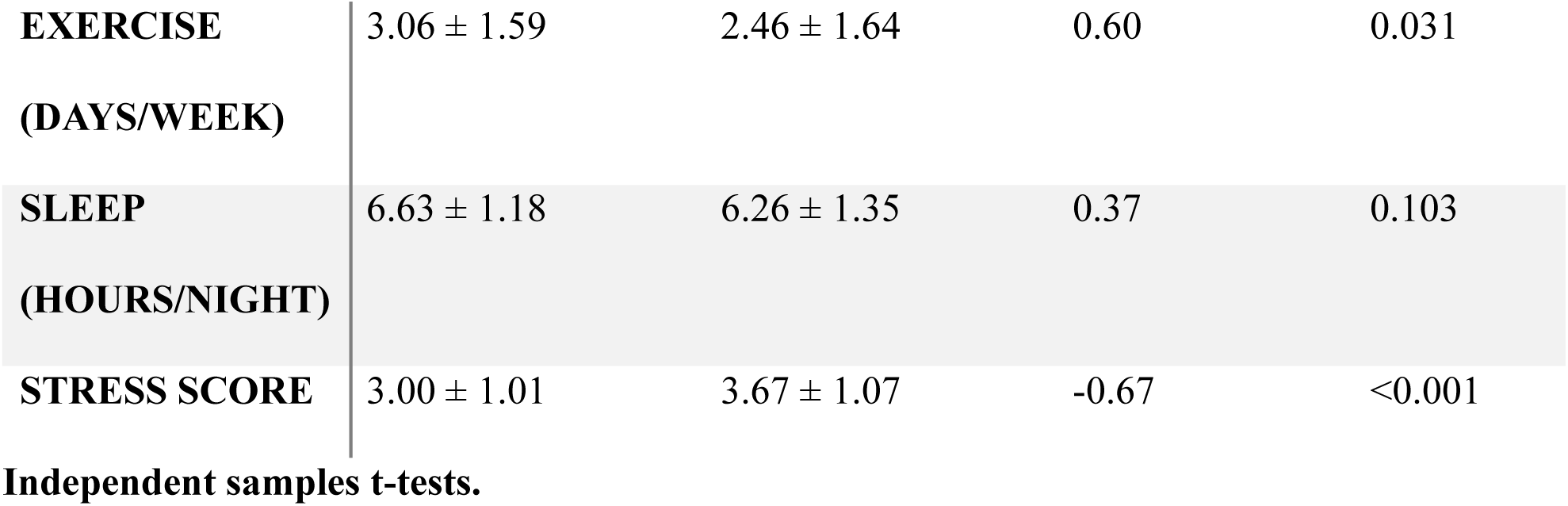
Lifestyle and Anthropometric Differences by Hypertension Status.

| VARIABLE | NO<br>HYPERTENSION<br>MEAN ± SD | HYPERTENSION<br>MEAN ± SD | MEAN<br>DIFFERENCE | P-VALUE |
| --- | --- | --- | --- | --- |
| **BMI** | $24.39 \pm 2.70$ | $28.19 \pm 3.43$ | -3.80 | <0.001 |
| **EXERCISE**<br>**(DAYS/WEEK)** | 3.06 ± 1.59 | 2.46 ± 1.64 | 0.60 | 0.031 |
| **SLEEP**<br>**(HOURS/NIGHT)** | 6.63 ± 1.18 | 6.26 ± 1.35 | 0.37 | 0.103 |
| **STRESS SCORE** | 3.00 ± 1.01 | 3.67 ± 1.07 | -0.67 | <0.001 |

Independent samples t-tests.

Significant regional disparities in hypertension prevalence were observed. Among participants from the Bay Area, **71.0% met criteria for hypertension**, compared with **97.9% of participants from the Central Valley** (χ²(2) = 31.2, p < 0.001).

Regional differences were even more pronounced for **undiagnosed hypertension**. In the Bay Area, **37.0% of participants exhibited previously undiagnosed hypertension**, whereas **91.7% of Central Valley participants had undiagnosed hypertension** (χ²(2) = 71.0, p < 0.001). Participants from other regions also demonstrated high rates of hypertension and undiagnosed hypertension, although these estimates should be interpreted cautiously due to small sample sizes.

**Table 4.**
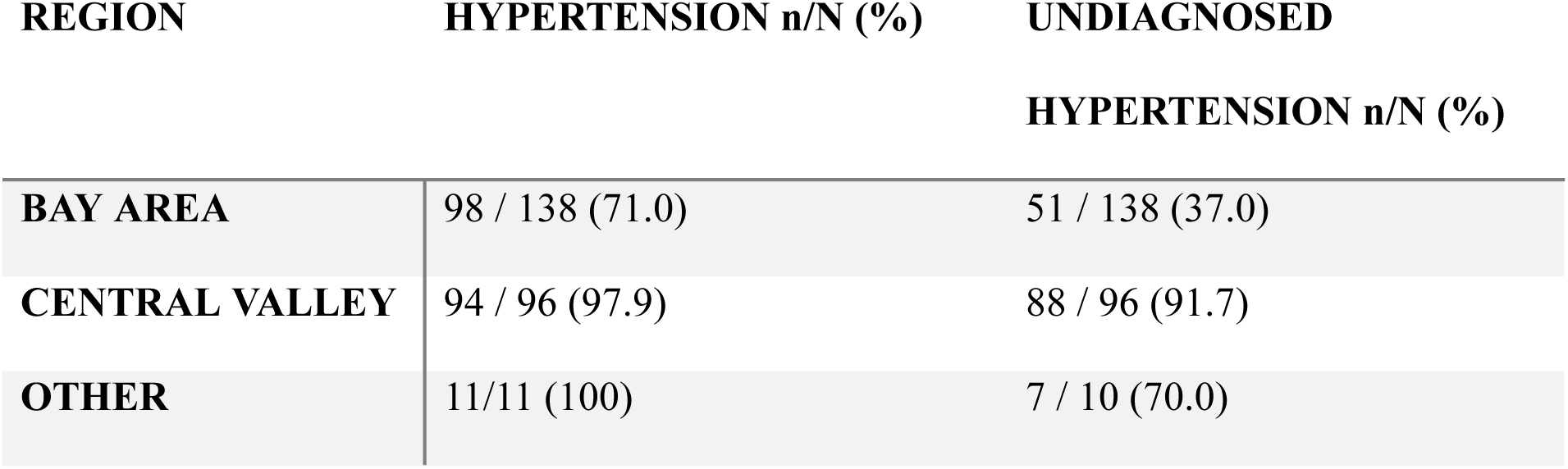
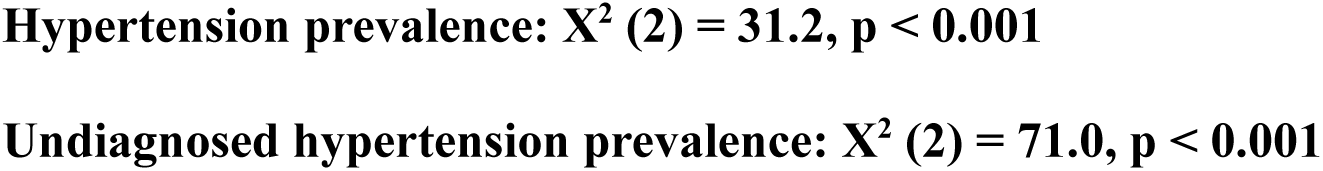
Regional Differences in Hypertension and Undiagnosed Hypertension.

To further evaluate factors associated with hypertension, a multivariable logistic regression model was performed adjusting for region, age group, gender, and BMI. Participants in the **Central Valley had significantly higher odds of hypertension compared with those in the Bay Area (OR = 14.29, 95% CI 2.89–70.63, p = 0.001)**. Higher BMI was also independently associated with hypertension, with each one-unit increase in BMI associated with a **26% increase in the odds of hypertension** (OR = 1.26, 95% CI 1.10–1.43, p < 0.001). Participants aged **25–34 years had higher odds of hypertension compared with those aged 18–24 years (OR = 5.76, 95% CI 1.01–32.98, p = 0.049)**, while other age groups were not statistically significant. Male participants had higher odds of hypertension compared with females, although this association did not reach statistical significance (OR = 2.24, 95% CI 0.98–5.14, p = 0.057).

**Table 5.** Multivariable Logistic Regression Predicting Hypertension.

| PREDICTOR | ODDS RATIO | 95% CI | p-VALUE |
| --- | --- | --- | --- |
| CENTRAL VALLEY<br>VS BAY AREA | 14.29 | 2.89 – 70.63 | 0.001 |
| AGE 25-34 VS 18-24 | 5.76 | 1.01 – 32.98 | 0.049 |
| AGE 35-44 VS 18-24 | 2.74 | 0.65 – 11.66 | 0.172 |
| AGE 45-54 VS 18-24 | 2.95 | 0.77 – 11.37 | 0.115 |
| AGE 55-64 VS 18-24 | 3.50 | 0.72 – 16.99 | 0.120 |
| AGE $\geq 65$ VS 18-24 | 0.79 | 0.03 – 19.91 | 0.885 |
| MALE VS FEMALE | 2.24 | 0.98 – 5.14 | 0.057 |
| **BMI** | 1.26 | 1.10 – 1.43 | <0.001 |

Footnote: The Other vs Bay Area contrast was not reported because all participants in the Other region with complete hypertension data were classified as hypertensive, resulting in complete separation and an unstable maximum-likelihood estimate. The regression model included 241 participants with complete data for all model variables.

Multivariable logistic regression analysis identified geographic region and BMI as significant predictors of hypertension. Participants in the Central Valley had higher odds of hypertension compared with those in the Bay Area after adjustment for age group, gender, and BMI.

## Discussion

This study examined the prevalence of hypertension and other cardiovascular risk factors among South Asian adults participating in community health screenings across Northern California. Several important findings emerged. First, hypertension was highly prevalent in this screened sample, with over 80% of participants with sufficient blood pressure data meeting criteria for hypertension. Second, a substantial proportion of participants exhibited previously undiagnosed hypertension, suggesting that a large segment of this population may have elevated cardiovascular risk without awareness or treatment. Third, significant regional disparities were observed: participants from the Central Valley had markedly higher rates of both hypertension and undiagnosed hypertension compared with participants from the Bay Area. Finally, individuals with hypertension demonstrated significantly higher BMI and stress levels and reported lower exercise frequency compared with participants without hypertension, while sleep duration did not differ significantly between groups.^20, 21^

The high prevalence of hypertension observed in this study is consistent with prior research showing that South Asians develop hypertension and other cardiometabolic diseases at younger ages and lower BMI levels than many other populations.^4, 22^ Genetic susceptibility, dietary patterns, and increased visceral adiposity may contribute to this elevated risk.^23^

One of the most striking findings of this study was the high prevalence of undiagnosed hypertension, particularly among participants from the Central Valley. While over one-third of Bay Area participants exhibited previously undiagnosed hypertension, more than 90% of Central Valley participants with elevated blood pressure reported no prior diagnosis. Differences in healthcare access, preventive screening availability, and primary care utilization may contribute to this regional disparity.^24^ The Central Valley has historically faced shortages of healthcare providers and reduced access to preventive services compared with urban regions of California.^25–27^ Community members in these areas may therefore be less likely to receive routine blood pressure screening or ongoing primary care, leading to delayed diagnosis of hypertension. These findings suggest that a substantial portion of cardiovascular risk in this population may remain clinically unrecognized. Multivariable analysis further indicated that geographic region and BMI were significantly associated with hypertension in this cohort. Participants from the Central Valley had substantially higher odds of hypertension compared with those from the Bay Area after adjustment for age group, gender, and BMI (OR = 14.29, 95% CI 2.89–70.63). The wide confidence interval, however, indicates substantial uncertainty in the magnitude of this association and warrants cautious interpretation.^28^

These findings highlight the potential value of community-based screening initiatives, particularly in underserved regions. Screening events conducted in culturally familiar settings such as religious institutions and community centers may help identify individuals with elevated cardiovascular risk who have limited engagement with the healthcare system.^29–31^ Early identification of hypertension is critical because timely lifestyle and pharmacologic interventions can substantially reduce cardiovascular morbidity.^32^

This study has several limitations. First, some blood pressure values were self-reported rather than directly measured during the screening event, which may introduce reporting bias.^33^ Second, blood pressure values were collected in categorical ranges for some participants, requiring estimation methods that may reduce measurement precision.^34^ Third, sufficient blood pressure data for hypertension classification were unavailable for some participants, resulting in a reduced sample size for hypertension-specific analyses. Fourth, the study relied on a convenience sample of individuals attending community screenings, which may not be representative of the broader South Asian population in Northern California and may overrepresent individuals with existing health concerns or elevated cardiovascular risk. Fifth, the relatively small number of participants without hypertension in the Central Valley resulted in imprecise estimates of the magnitude of the regional association, as reflected by the wide confidence interval in the multivariable model. Sixth, the cross-sectional design limits the ability to infer causal relationships between lifestyle factors, regional characteristics, and hypertension status.^18^ Finally, while the study captured regional differences between the Bay Area and Central Valley, additional research is needed to further explore the structural, healthcare access, and other contextual factors contributing to these disparities.

Despite these limitations, this study identifies substantial opportunities to improve cardiovascular disease prevention among South Asian communities in Northern California. Future efforts should focus on expanding culturally tailored screening and prevention programs, particularly in underserved regions where access to diagnosis and routine preventive care may be limited.^35, 36^

## Data Availability

The de-identified data supporting the findings of this study are available from the corresponding author upon reasonable request, subject to applicable ethical, privacy, and institutional requirements.

